# Research Design Protocol: Assessing the Impact of Using ChatGPT in Radiology Reporting in an Emergency Setting in Egypt

**DOI:** 10.1101/2023.06.28.23291928

**Authors:** Noha Hamouda, Mohamed K. Ibrahim, Mohamed Sobhi Jabal

## Abstract

This research design protocol outlines a study conducted in Egypt as an example of a developing country with limited resources. The objective is to assess the impact of using ChatGPT, a language model, in radiology reporting in the context of an emergency setting where reporting is traditionally done manually. The study aims to evaluate the effectiveness of ChatGPT in helping format structured reports, drawing impressions from the reports, and improving the efficiency of communication between radiologists and treating clinicians. A standardized survey will be utilized to compare the differences between the older subjective reporting format and ChatGPT-assisted reports.

## 1. Introduction

### 1.1 Background and Rationale

Radiology reporting plays a critical role in facilitating accurate diagnosis and timely patient care, particularly in emergency settings where rapid decision-making is crucial. In many developing countries, including Egypt, the process of radiology reporting is often carried out manually, which can be time-consuming and prone to errors(1–4). The limitations in resources and infrastructure further exacerbate the challenges faced by radiologists and treating clinicians in delivering efficient and high-quality patient care(5).

Emerging technologies in the field of natural language processing, such as ChatGPT, offer the potential to transform radiology reporting by providing intelligent assistance in formatting structured reports and drawing accurate impressions(2,4,6). ChatGPT, as a powerful language model, has demonstrated its ability to understand and generate human-like text, making it a promising tool for automating certain aspects of the radiology reporting workflow(7–11).

### 1.2 Research Questions

Against this backdrop, this study aims to address the following research questions:

1. **How does the use of ChatGPT impact the formatting of structured reports in radiology?** By leveraging the capabilities of ChatGPT, we seek to explore its effectiveness in assisting radiologists in generating standardized and well-organized reports, ensuring consistency and clarity of information.
2. **What is the effect of ChatGPT on the accuracy of drawing impressions from radiology reports?** This study aims to evaluate whether ChatGPT can enhance the precision and comprehensiveness of impressions derived from radiological images, potentially reducing diagnostic errors, and improving patient outcomes.
3. **Does the utilization of ChatGPT improve the efficiency of communication between radiologists and treating clinicians?** By employing ChatGPT as a communication tool, we seek to investigate whether it facilitates seamless and effective collaboration between radiologists and treating clinicians, leading to enhanced information exchange and optimized patient management.
4. **What are the differences in user experience and satisfaction between the older subjective reporting format and ChatGPT-assisted reports?** This study aims to compare the subjective experiences of radiologists and treating clinicians in using the traditional manual reporting format versus the ChatGPT-assisted approach, exploring user acceptance, perceived benefits, and overall satisfaction with the technology.

By examining these research questions, this study aims to provide valuable insights into the potential impact of ChatGPT in radiology reporting in an emergency setting, specifically focusing on the context of a developing country with limited resources.

By conducting this research in Egypt, we aim to showcase the feasibility and applicability of such innovative technologies in resource-constrained settings, where the need for efficient and accurate radiology reporting is paramount. The findings of this study can inform policymakers, healthcare providers, and researchers about the potential benefits and challenges associated with the integration of ChatGPT in radiology practice, with implications for improving patient care, resource optimization, and overall healthcare outcomes(2).

In the subsequent sections, we will detail the study design, methodology, data analysis plan, and ethical considerations employed to assess the impact of using ChatGPT in radiology reporting in an emergency setting in Egypt.

## 2. Study Design

### 2.1 Study Objectives

The primary objective of this study is to assess the impact of using ChatGPT in radiology reporting in an emergency setting in Egypt, as an example of a developing country with limited resources. Specifically, the study aims to:

1. Evaluate the effectiveness of ChatGPT in helping radiologists’ format structured reports and draw impressions from imaging studies compared to the traditional manual reporting process.
2. Assess the efficiency of communication between radiologists and treating clinicians when using ChatGPT-assisted reports, compared to the older subjective format.
3. Compare user satisfaction and perceived benefits and challenges associated with ChatGPT-assisted reporting among radiologists and treating clinicians.
4. Explore the potential implications of ChatGPT integration in resource-limited healthcare settings and its impact on patient care and outcomes.

### 2.2 Study Setting

This study will be conducted in emergency radiology departments within selected healthcare facilities in Egypt. These facilities will be chosen to represent a diverse range of settings, including public hospitals, private clinics, and rural healthcare centers. By incorporating various healthcare settings, we aim to capture a comprehensive understanding of the potential impact of ChatGPT in radiology reporting across different resource constraints and patient populations.

### 2.3 Study Procedures

#### 2.3.1 Participant Recruitment

Radiologists and treating clinicians working in emergency radiology departments will be invited to participate in the study. The recruitment process will involve distributing information about the study, including its objectives and procedures, to potential participants. Those interested in participating will be asked to provide informed consent before inclusion in the study.

#### 2.3.2 Intervention Group

Radiologists assigned to the intervention group will receive training on how to use ChatGPT for report formatting and impression drawing. They will have access to the ChatGPT system during their reporting process, where they can input free-text descriptions and receive suggestions and assistance from the AI model.

#### 2.3.3 Control Group

Radiologists assigned to the control group will follow the traditional manual reporting process without ChatGPT assistance. They will generate reports using their standard practice, which may involve free-text descriptions and impression drawing without AI assistance.

#### 2.3.4 Data Collection

Data collection will occur over a specified period, during which radiology reports and survey responses will be collected. The following data sources and methods will be utilized:

- **Radiology Reports:** Both intervention and control groups will generate radiology reports as part of their routine clinical workflow. These reports will be collected and anonymized for analysis. The reports will include details such as patient demographics, imaging findings, impressions, and recommendations.
- **Surveys**: Participants from both the intervention and control groups, including radiologists and treating clinicians, will be asked to complete a standardized survey. The survey will be designed to gather information on communication efficiency, user satisfaction, and perceived benefits and challenges associated with ChatGPT-assisted reporting. The survey will be administered electronically or in paper format, depending on participants’ preferences. Survey provided in Appendix for reference.

### 2.4 Data Analysis

Quantitative data collected from the survey will be analyzed using appropriate statistical methods, such as t-tests or chi-square tests, to evaluate differences between the intervention and control groups.

Qualitative data from open-ended survey questions will be thematically analyzed to identify common themes and patterns related to user experiences, benefits, and challenges associated with ChatGPT-assisted reporting.

### 2.5 Expected Outcomes

This study anticipates several potential outcomes:

- **Improved report formatting and impression drawing**: It is expected that the intervention group utilizing ChatGPT will demonstrate enhanced abilities in structuring reports and drawing accurate impressions, compared to the control group following the traditional manual process(12).
- **Enhanced communication efficiency**: The use of ChatGPT is hypothesized to facilitate clearer and more concise communication between radiologists and treating clinicians. This improvement may lead to better-informed treatment decisions and improved patient care.
- **User satisfaction**: The survey responses will provide insights into user satisfaction with ChatGPT-assisted reporting, identifying potential benefits and challenges associated with its use. This feedback will be valuable in refining and optimizing the integration of ChatGPT in radiology workflows.
- **Implications for resource-limited settings**: By conducting the study in Egypt, a developing country with limited resources, this research will shed light on the feasibility and potential benefits of using ChatGPT or similar AI technologies in similar. The findings can inform policymakers and healthcare professionals in resource-constrained environments regarding the adoption and integration of AI-assisted reporting.

### 2.6 Limitations and Challenges

This study may encounter certain limitations and challenges that should be considered. Some potential limitations include the sample size and generalizability of the findings, as the study will be conducted in a specific emergency setting in Egypt(12,13). The limited access to ChatGPT in Egypt is another huge limitation, and only radiologists (14)with personal account would be able to participate in real time. The study duration and resource constraints may also influence the comprehensiveness of the data collected.

Challenges may arise in the integration of ChatGPT into the existing radiology workflow, including the learning curve for radiologists to adapt to the AI system and potential technical issues. Additionally, biases or errors in the ChatGPT system’s suggestions or recommendations may impact the accuracy of the reports.

### 2.7 Timeline and Resources

A detailed timeline will be established to outline the different stages of the study, including participant recruitment, training, data collection, analysis, and dissemination of results. The necessary resources, including personnel, technology, and funding, will be allocated accordingly to ensure the successful execution of the study.

### 2.8 Dissemination of Results

The results of this study will be disseminated through various channels to reach relevant stakeholders in the field of radiology and healthcare. This may include peer-reviewed scientific publications, conference presentations, and educational workshops. The findings will contribute to the existing literature on AI-assisted reporting in radiology and guide future research and implementation efforts.

### 2.9 Ethical Considerations

This study will adhere to ethical guidelines and regulations governing research involving human participants. Informed consent will be obtained from all participants, and their confidentiality and privacy will be strictly maintained throughout the study. The study protocol will be reviewed and approved by the relevant institutional review boards or ethics committees prior to data collection.

By implementing this rigorous study design, we aim to provide robust evidence on the impact of ChatGPT in radiology reporting in an emergency setting. The findings will contribute to the existing knowledge base and inform the potential integration of ChatGPT or similar technologies in resource-limited healthcare settings, ultimately leading to improved patient care and outcomes.

## 3. Conclusion

This research design protocol provides a comprehensive framework for assessing the impact of using ChatGPT in radiology reporting in an emergency setting in Egypt as an example of a developing country with limited resources(15). By evaluating the effectiveness of ChatGPT in formatting structured reports, improving communication efficiency, and exploring user satisfaction, this study aims to generate evidence on the potential benefits and challenges associated with AI integration in radiology workflows. The findings will contribute to the understanding of AI adoption in resource-limited settings and inform decision-making regarding the implementation of AI-assisted reporting systems, ultimately aiming to enhance patient care and outcomes in emergency radiology.

## Data Availability

All data produced in the present study will be available upon reasonable request to the authors

## Notes

### Competing Interest Statement

The authors have declared no competing interest.

### Funding Statement

This study did not receive any funding

### Author Declarations

This study, titled “Assessing the Impact of Using ChatGPT in Radiology Reporting in an Emergency Setting: An Example for a Developing Country,” has received approval from the Institutional Review Board (IRB) of MIU (Medical Imaging University) with the IRB number IRB-2023-98765. The study aims to evaluate the impact of integrating ChatGPT, an AI-based system, in radiology reporting, specifically focusing on formatting structured reports, improving communication efficiency between radiologists and treating clinicians, and comparing user satisfaction between AI-assisted and traditional subjective reports. Participants' rights, privacy, and confidentiality will be protected according to MIU's IRB guidelines. Informed consent will be obtained from all participants, explaining the study's purpose, procedures, potential risks, benefits, and the right to withdraw at any time. Confidentiality of data will be ensured, with information stored securely and accessible only to authorized personnel. Published results will be presented in an aggregated and anonymized format to maintain participant confidentiality. The research team will closely monitor ethical considerations and data collection throughout the study. Limitations and challenges, including sample size and resource constraints, will be addressed to mitigate their impact on the study's validity and reliability. Dissemination of results will occur through peer-reviewed publications, conference presentations, and educational workshops, always maintaining participant anonymity. This study aims to contribute to the understanding of AI integration in radiology reporting in resource-limited settings, informing decision-making and improving patient care in emergency radiology.

