## Appendix for "Research Design Protocol: Assessing the Impact of Using ChatGPT in Radiology Reporting in an Emergency Setting in Egypt"

### Tools:

In this study, we will use two assessment tools to evaluate the impact of using ChatGPT in radiology reporting in the emergency setting: a standardized checklist for report formatting assessment and a structured survey to measure the efficiency of communication between radiologists and clinicians.

#### 1. Standardized Checklist for Report Formatting Assessment:

To assess the impact of ChatGPT assistance on report formatting, we developed a standardized checklist that evaluated specific aspects of the radiology reports. The checklist included the following formatting aspects:

- **Structured Organization:** This aspect assessed the presence and appropriate organization of key sections in the radiology report, such as patient demographics, clinical history, technique, findings, and impressions.
- **Completeness of Content**: This aspect examined the inclusion of all relevant findings, impressions, and recommendations in the report.
- **Clear Findings Section:** This aspect evaluated the clarity and organization of the findings section, ensuring that the radiologist's observations were well presented and easily understandable.

For each aspect, reports will be evaluated as meeting or not meeting the specified formatting criteria. The checklist provides a quantitative measure of the improvements in report formatting achieved with ChatGPT assistance.

#### 2. Structured Survey for Communication Efficiency:

To assess the efficiency of communication between radiologists and clinicians, we designed a structured survey that focused on the clinicians' perception of the radiology reports. The survey included Likert scale questions that measured the clarity, comprehensibility, and ease of understanding of the reports. The Likert scale ranged from 1 (poor) to 5 (excellent), allowing respondents to rate their experience with both manual and ChatGPT-assisted reports.

In addition to Likert scale questions, the survey also included open-ended questions to gather qualitative feedback from the clinicians. These questions aimed to explore their opinions on the advantages and challenges of using ChatGPT-assisted reports in their clinical practice.

The structured survey provided quantitative data to compare the communication efficiency of manual reports versus ChatGPT-assisted reports. It also allows clinicians to provide valuable qualitative insights into their experiences with the new reporting system.

By employing these assessment tools, we will be able to gather comprehensive data on the impact of ChatGPT in radiology reporting, both in terms of objective report formatting improvements and subjective perceptions of communication efficiency.

### Example report:

**Example of a Manual Report:**

**Patient**: ####

**Age**: 45

**Gender**: Male

**Study**: Chest X-ray

**Date**: June 20, 2023

**Findings:**

The chest X-ray shows consolidation in the right lower lobe, suggestive of pneumonia. No other significant abnormalities are observed.

**Impression:**

Right lower lobe pneumonia.

**Example of a ChatGPT-Assisted Report:**

**Patient**: ####

**Age**: 45

**Gender**: Male

**Study**: Chest X-ray

**Date**: June 20, 2023

**Findings**:

The chest X-ray reveals consolidation in the right lower lobe, with air bronchograms and loss of volume. The consolidation appears to be of bacterial origin, consistent with pneumonia. No evidence of pleural effusion or mediastinal abnormalities is noted.

**Impression**:

1. Right lower lobe pneumonia with typical radiographic features.

2. No associated complications such as pleural effusion or mediastinal abnormalities identified.

**Comparison:**

In the manual report, the findings are described briefly, mentioning the presence of consolidation in the right lower lobe indicative of pneumonia. The impression provides a concise conclusion stating "Right lower lobe pneumonia."

On the other hand, the ChatGPT-assisted report offers more comprehensive details in the findings section. It describes the consolidation as being accompanied by air bronchograms and loss of volume, suggesting bacterial etiology. Additionally, it explicitly mentions the absence of pleural effusion or mediastinal abnormalities. The impression is also more comprehensive, providing a detailed description of the pneumonia findings and emphasizing the absence of associated complications.

The ChatGPT-assisted report, with its structured and detailed format, offers a more comprehensive and informative assessment of the radiological findings. It provides a clearer understanding of the extent and characteristics of pneumonia, aiding in clinical decision-making and facilitating effective communication between radiologists and clinicians.

### Survey:

**Translation of the survey questions (Original in Arabic):**

**Survey for Evaluating Communication Efficiency in Radiology Reports using ChatGPT.**

**Part 1: Demographic Information**

1. Name: _______________________________

2. Role:

- Radiologist

- Clinician (Please specify your specialty: ________________)

**Part 2: Perception of Radiology Reports:**

Please rate the following statements based on your experience with both manual and ChatGPT-assisted radiology reports. Use the Likert scale below, where 1 indicates "Strongly Disagree" and 5 indicates "Strongly Agree."

**1. Clarity of the reports:**

Clarity of the reports:

- Manual Reports: 1 2 3 4 5

- ChatGPT-Assisted Reports: 1 2 3 4 5

**2. Ease of understanding:**

Comprehensibility of the reports:

- Manual Reports: 1 2 3 4 5

- ChatGPT-Assisted Reports: 1 2 3 4 5

**3. Completeness of information:**

How do you rate the completeness of information in and ease of understanding the findings and impressions:

- Manual Reports: 1 2 3 4 5

- ChatGPT-Assisted Reports: 1 2 3 4 5

**4. Structural organization of the reports**

- Manual Reports: 1 2 3 4 5

- ChatGPT-Assisted Reports: 1 2 3 4 5


**Part 3: Open-Ended Questions**

Please provide your opinion on the advantages and challenges of using ChatGPT-assisted reports compared to manual reports. Your insights are highly appreciated.

1. What do you perceive as the advantages of ChatGPT-assisted reports?

2. What challenges, if any, did you encounter when using ChatGPT-assisted reports?

3. Do you have any suggestions for further improvement or customization of the ChatGPT system for radiology reporting?

**Part 4: Additional Comments**

If you have any additional comments or feedback regarding the impact of ChatGPT on radiology reporting, please share them here.

Thank you for your participation in this survey. Your input is crucial in helping us evaluate the impact of ChatGPT on radiology reporting and improve the communication between radiologists and clinicians.
